# Mobilisation towards formal employment in the healthcare system: A qualitative study of community health workers in South Africa

**DOI:** 10.1101/2023.07.11.23292526

**Authors:** Hlologelo Malatjif, Frances Griffiths, Jane Goudge

## Abstract

In low and middle-income countries (LMICs), community health workers (CHWs) play a critical role in delivering primary health care (PHC) services to vulnerable populations. In these settings, they often receive low stipends, function with a lack of basic resources and have little bargaining power with which to demand better working conditions. In this article, we examine CHWs’ employment status, their struggle for recognition as health workers, and their activities to establish labour representation in South Africa. Using a case study approach, we studied seven CHW teams located in semi-urban and rural areas of Gauteng and Mpumalanga Provinces, South Africa. We used in-depth interviews, focus group discussions and observations to gather data from CHWs and their representatives, supervisors and PHC facility staff members. The rural and semi-urban sites CHWs were poorly supervised, resourced and received meagre remuneration, their employment outsourced, without employment benefits and protection. The lack of career progression opportunities demotivated the CHWs, particularly those keen to establish a career in health. In the semi-urban sites, CHWs established a task team to represent them that held regular meetings and often used violent and disruptive strategies against clinic, district and provincial management, which often led to tensions and conflicts with facility staff and programme coordinators. After a meeting with the local provincial legislature, the task team joined a labour union (NEHAWU) in order to be able to participate in the local Bargaining Council. Though they were not successful in getting the government to provide permanent employment, the union negotiated an increase in stipend from R2 500 (136 USD) to R3 500 (192 USD). In contrast, in the rural sites, the CHWs were not actively demanding permanent employment due to their employment contracts being partly managed by non-government organisations (NGOs); they were fearful of being recalled from the government programme. After the study ended, during the height of COVID-19 in 2020, when the need for motivated and effective CHWs became much more obvious to decision makers, the semi-urban-based teams received permanent employment with remuneration between R9-11,000 (500-600 USD). The task team and their protests raised awareness of the plight of the CHWs, and joining a formal union enabled them to negotiate a modest salary increase. However, it was the emergency created by the world-wide COVID-19 pandemic that forced decision-makers to acknowledge their reliance on this community-based cadre. Hopefully this recognition, and the associated gains, will not fade as the pandemic recedes.

## INTRODUCTION

In pursuit of universal health coverage (UHC), low and middle-income countries (LMICs) are investing in programmes to extend primary health care services (PHC) to marginalised communities [1]. Community health worker (CHW) programmes are being revitalised to extend the delivery of the PHC services to marginalised communities and strengthen the provision of community-based health services [2]. Systematic reviews have shown CHWs are able to extend access to PHC services by supporting chronic care management, antenatal and postnatal support programmes, and early identification of malnourished children [2–4].

However, in some settings the programmes fail to deliver on expectations due to limited access to resources (e.g. blood pressure and glucose monitoring machines), low and sometimes irregular remuneration and poor integration into the healthcare system resulting in poor relationships with facility-based staff members [3, 5, 6]. CHW are mostly women, often with low levels of education, who are expected to work on voluntary basis, or who are underpaid with no permanent employment security [7]. These employment conditions contribute to CHWs’ low work morale, demotivation and high work turnover [1, 8]. Pandya et al. argues the provision of incentives, both monetary and non-monetary is crucial to improve CHW motivation, job satisfaction, and performance, which, in turn, improves retention [7]. Monetary incentives include stable salaries, while non-monetary incentives include the provision of working tools such as uniforms and raincoats.

While the World Health Organisation (WHO) has argued for the optimisation of CHW programmes through fair remuneration schemes and provision of career development pathways, as well as integration into the health system and provision of resources [6], there has been little progress. Unionisation among the CHWs is often low due to CHWs working in isolation or in pairs, with little bargaining power with which to demand better working conditions [15, 20].

In South Africa labour unions have a played a key role in the country’s history in opposing Apartheid; the white minority government had enacted labour laws based on race categorisation thus resulting in unfair labour practices [9]. In the 1990s, the Congress of South African Trade Unions (COSATU), an umbrella body of unions, formed an alliance with the governing political party – the African National Congress. As a result, unions participated in government policy discussions, and influenced labour legislation strengthening employees’ rights in workplace. Public sector unions’ alliance with government enabled the formally employed workers to consistently obtain above inflation salary increases through centralised bargaining, although union leaders are often criticized for using the alliance to build their own political connections, rather than representing the concerns of ordinary workers [10, 11].

In line with international UHC goals, in 2011, the South African Department of Health (DOH) has attempted to strengthen the delivery of PHC services [12]. A nation-wide CHW programme (locally known as ward-based outreach teams; WBOT) was introduced. The programme intention was for each WBOT, linked to a local PHC facility, to comprise of 6 CHWs who serve a defined geographic area, provide a combination of promotive and preventative healthcare services to households, and make referrals to social services workers. The team was to be led by a nurse and supported by a health promoter and an environmental officer.

Prior to the introduction of the WBOTs, the CHWs were in the employment of non-government organisations (NGOs), mainly providing HIV/AIDS and TB related services at community level [13]. The NGO programmes were fragmented and single disease-focused, thus neglecting the other needs of individuals and families. The WBOTs replaced the NGO-led programmes and absorbed the CHWs on a 12-month renewable contract earning a monthly stipend of R2 500 (136 USD) (at the time of study), their employment was outsourced to either NGOs or payroll administration company. As a result, despite their employment within a nationwide CHW programme, the CHWs had little employment security and were paid a meagre stipend. With high levels of union activity being common in South Africa, informal groupings of CHW have been protesting for several years, demanding better working conditions [19, 20, 26]. In this article, we examine CHWs’ employment status and their mobilisation for formalisation in the healthcare system.

## METHODS

### Study design

The data originate from a large project and a doctoral study that investigated the design and implementation of CHW programmes in South Africa. The larger project included the introduction of a nurse mentor intervention with a specific focus on building CHWs knowledge and skills, supporting their integration into healthcare systems and community structures. Together, the two studies adopted a qualitative approach with 7 case studies (each case study being a team of CHWs).

### Study setting

The research was carried out in Sedibeng, Johannesburg and Tshwane health districts of Gauteng Province and Ehlanzeni health district of Mpumalanga Province. The two provinces were selected based on their location (urban vs rural), history and contrasting models of implementing the CHW programmes. The CHW programmes in the three districts of Gauteng province were implemented by the Provincial Department of Health with a payroll administration company contracted to pay the CHWs on behalf of the department. The Mpumalanga province, where the rural case studies were located, had not fully implemented the national CHW programme. Here, the Provincial Department of Health funded existing local NGOs, that provided home-based care services, to deploy some of its carers to constitute the CHW programme in local health facilities. The NGOs remained the primary employer of the CHWs, while supervision and resources were being provided by the health facilities.

### Data collection

The data was collected by the first author and a group of data collectors from 2016 to 2019. The first author was a South African Research Chairs Initiative (SARChI) doctoral researcher employed by a research centre undertaking health policy and systems research. The data collectors were qualified qualitative researchers sensitised to the type of activities CHWs undertake. Individual interviews, focus group discussions and observations were used to collect data from the different CHW teams in the two provinces. The research instruments were all in English, during interviews the data collectors translated the interview questions to local languages (i.e. IsiZulu, Sesotho and Sepulana) for participants who could not understand the English language. To ensure the consistency of the translations, data collectors participated in role-plays to practice posing the same question in the different languages prior to field work.

#### Focus group discussions

Nine focus group discussions (FGDs) were held in the different sites, and each FGD had approximately 10 participants, all women (Table 1). All CHWs who formed part of the CHW team in the health facilities were invited to participate in FGDs, and there were no reported refusals. We used a structured guide to ask the CHWs about their working conditions, daily activities, resources they require to perform their duties and their engagement with the facility and Department of Health. Each of the discussions were audio-recorded and lasted approximately 2 hours. The first author facilitated the discussions while the data collectors took notes. The notes were typed by the data collectors and formed part of the dataset for analysis.

**Table 1:** Data collection methods and number of participants

|  | Sedibeng |  | JHB | Tshwane |  | Ehlanzeni |  | Total |
| --- | --- | --- | --- | --- | --- | --- | --- | --- |
|  | Team 1 | Team 2 | Team 3 | Team 4 | Team 5 | Team 6 | Team 7 |  |
| Focus group discussions with CHWs | 1 | 1 | 2 | 2 | 2 | 1 | 1 | 9 |
| Interviews with CHW teams | 4 | 4 | 3 | 2 | 1 | 1 | 2 | 17 |
| Interviews with nurse mentor | 4 |  | - | - |  | - |  | 4 |
| Interviews with task team members/<br>CHW representatives | 6 |  | - | - | - | - | - | 6 |
| Interviews with key informants | 5 |  | 2 | 6 |  | 3 |  | 16 |
| Observations of task team meetings | 6 (meetings) |  | - | - |  | - |  | 6 |

#### Interviews

We purposively selected and interviewed CHWs and their supervisors, facility staff members, nurse mentor and programme coordinators at district level. We asked them about the CHW programme (i.e. CHWs duties in households, resources, access to supervision, and conditions of employment). Furthermore, in Sedibeng district where we spent prolonged time in the field, we interviewed CHWs who represented their colleagues at district level meetings and when there were conflicts. The CHW representatives were referred to as a task team. We asked them about the origin of the task team, engagement with health facilities, district and province, and successes and challenges in advocating for CHWs rights. Each interview lasted approximately 45 minutes and were audio-recorded with participant consent.

#### Observations

These were conducted in order to understand the CHW activities relating to labour mobilisation aimed at challenging the employer to provide better conditions of employment. We observed 6 CHW representatives meetings with the general CHW population, programme coordinators and the payroll administration company (Table 1). In these meetings, we documented the issues being discussed and resolutions made.

### Data analysis

As described by Braun [14], we used thematic content analysis method to analyse the data. The team read through the interview transcripts and meeting notes and developed a codebook. Codes captured information about CHW resources requirement (e.g., what they have and do not have) and activism to challenge the government to provide permanent employment. The research team held regular meetings to discuss the coded data and emerging themes. We undertook comparative analysis between the different cases of context, processes and outcomes of mobilisation.

### Ethics approval and participant consent

The larger project was cleared by the University of the Witwatersrand HREC Medical Committee (M160354) and the Sedibeng health district. The doctoral study received ethical clearance (M180540) from the same university ethics body, and the Johannesburg, Tshwane and Ehlanzeni provincial government research authorities. The participants provided written informed consent before participation in interviews and focus groups. Before observing CHW in the field and their representative meetings with district management, we checked with those present for verbal consent. We explained there would be no audio-recording made; the researcher will write notes of the deliberations. There were no reported refusals in the study.

## FINDINGS

The CHWs conditions of employment and mobilisation to challenge government to provide permanent employment and resources is presented in two sections. The first section focuses on CHWs conditions of employment (i.e. stipend, employment security, career advancement opportunities, work tools and access to supervision), and how these conditions hindered the effectiveness of the CHWs in the different sites. The second part considers CHW labour mobilisation towards permanent employment.

### CHW conditions of employment

#### Stipend

Across the sites, the CHWs said that the low stipend was insufficient and demotivating: “…*the government should hear us and increase our stipend, so when we go to the community to motivate and counsel people, they understand us. We cannot talk to people who are frustrated and hungry when ourselves are also frustrated and hungry*.” (CHW, FGD 1, Tshwane District Team 2). The CHWs felt undervalued: “*We become de-motivated because of the stipend. Our stipend is too little, and you see the amount of work that we do. We get burned by the sun*” (CHW, FGD, Tshwane District Team 2). The CHWs provided services to patients located in informal settlements, hostels and remote areas. Some of the areas, particularly the hostels and informal settlements were unsafe due to high crime rates.

In the urban sites, the department contracted a payroll administration company to pay the CHWs (Table 2). The company issued a bank card that the CHWs used to access their stipend, however, the card did not have features of a traditional bank card (e.g., cash deposit and transfer options); it was only for the purpose of receiving and withdrawing the stipend. The company offices were located far from the reach of many CHWs, who lived and worked in the periphery of the districts, making it difficult to resolve any problems with the card or the stipend.

**Table 2:** Description of CHW programmes and labour representation

|  | Sedibeng (semi-urban) | Johannesburg (semi-urban) | Tshwane (semi-urban) | Ehlanzeni (rural) |
| --- | --- | --- | --- | --- |
| **Programme management** | Government |  |  | Government; contracted to local NGOs |
| **Salaries disbursement** | Paid by a payroll administration company contracted by the government |  |  | Paid by NGOs contracted by government; delays in disbursing salaries |
| **Member of union/ advocacy group** | Yes; prior to joining a formal labour union CHWs were represented by a task team | Yes | Yes | No |
| **Labour mobilisation activities** | Regular protests to demand permanent employment<br>Facility stay aways | Protests to demand permanent employment | Protests to demand permanent employment | None |
| **Outcomes of mobilisation** | Monthly stipend increased<br>Payroll administration company contract not renewed<br>In Sedibeng, the task team negotiated with facilities to attend to CHWs needs (e.g. resources) |  |  | Monthly stipend increased |

In the rural sites, where the programme was managed by local NGOs, the CHWs regularly received the stipend late, and would continue working for up to 6 months without being paid: “*Normally we are getting paid after 3 or 6 months so now we might get paid in October/ November. This is disturbing us. We are always in debt and when we get that amount even if it is back pay it all goes to the creditors*” (CHW, FGD, Ehlanzeni District Team 2). The sub-district officials were aware of the delayed payment of CHW stipend and expressed how it is important for the CHWs to be formal employees of the department, so they can be paid on time: “*The CHWs receive their stipends from the home-based care organizations (NGOs)*.

*The minister mentioned that the CHWs should be part of the department and have PERSAL numbers but due to the current [budget] constraints, I don’t see this happening soon.*” (Government official, Interview, Ehlanzeni District). The official blamed the NGOs as they always submit paperwork required by the department late for timely processing and release of funds.

#### Employment security

The urban-based teams had to renew their contracts with the payroll administration company every year, and this created anxiety among the CHWs: “*What if they wake up and say that they are not renewing our contracts? What are we going to do? We have children and families, the children are waiting on us as their moms to bring them something*” (CHW, FGD, Johannesburg District). The CHWs also felt the company, as contractor to the department, was disinterested in their call for permanent employment: “*I can tell you that payroll administration company doesn’t care about us. If our task team says that it has a meeting, the [payroll company] does not understand or give us the go ahead to attend the meeting. We have to struggle to go to that meeting.*” (CHW, FGD, Sedibeng District Team 1).

The CHW in rural sites were anxious about their contractual arrangement with the NGOs, as their employment was decided by the NGO managers. During our discussions, they seemed unable to openly discuss their employment concerns, as they feared being recalled from the government programme (where there was a possibility of permanent employment) by the NGO managers. These concerns often emerged after audio-recording had stopped.

#### Career advancement opportunities

Some CHWs were frustrated by the lack of career progression in the field, as they wanted to establish a career in health care (e.g., to be trained as a nurse). The Johannesburg team had young CHWs with matric qualifications who were interested in furthering their studies. They felt the Department of Health was not making opportunities available for them: “*In our team, there are people with mathematics and physical science; why can’t they take these people so that they could study for something like nursing, pharmacy and so forth? They can see that we love what we are doing. Not all of us have the money to go to private institutions to study. The government is failing us”* (CHW, FGD, Johannesburg District). One of the CHWs was already studying towards a teaching qualification at a local university. This was not true for the other sites, where the majority of the CHWs did not possess a matric pass, which is a prerequisite to enrol for tertiary education. However, there was still a desire among the CHWs to be trained and promoted into better positions with benefits: “*We complete trainings. At least when you have done the training, they should promote you but now we remain the same.*” (CHW, FGD, Johannesburg). A representative of the CHWs in Sedibeng was concerned that the Department of Health provides short training courses but never considers them for promotion after the training: “*After some of them finished and got their certificates; they are still doing the same work, same level, same stipend. So that is our biggest challenge. Why are there trainings that don’t have opportunities?*” (CHW representative, Interview 1, Sedibeng District).

#### Work tools

When undertaking household visits, CHWs require resources such as blood pressure and glucose monitoring machines, stationary and a raincoat etc. Across the different sites, the majority of the CHWs did not have these resources. At the inception of the programme, the employer made provisions for the CHWs to have these resources, although some teams did not receive them: “*We only saw the boxes. One sister from clinic X did ask us how come we don’t have BP machines. We said we don’t know. Then the sister asked the sister in-charge and she promised to get us BP machines. We got 2 machines and we had to share them but now there are no batteries*.” (CHW, FGD, Ehlanzeni District Team 2).

The lack of equipment affected the CHW activities in the households: “*Some patients expect us to check their blood pressure and sugar level when we visit them. If I don’t have the machines, they ask what the use of me visiting them?*” (CHW, FGD, Ehlanzeni 6). Another CHW commented: “*The patients keep on asking us to measure their BP and blood sugar…we always tell them that we don’t have equipment. That makes people to undermine us; they say we are useless*.” (CHW, FGD, Johannesburg District).

A facility manager in Tshwane was aware the CHWs needed resources, however, he felt helpless: “*They received backpacks, but they did not have all the equipment. The CHWs are supposed to have blood sugar machines but they don’t have them. If the clinic were to distribute the glucose strips to all the CHWs, the clinic will be left with none and that wouldn’t be good because they are very costly.*” (Facility Manager, Interview, Tshwane District Team 4).

Space within the health facilities was an issue in both urban and rural sites. The CHWs in Johannesburg did not have a dedicated space to use to meet, discuss work and store patient records. A CHW commented: “… *sometimes we find that our things are missing or when you check your stuff you will find your things rearranged as if someone was looking for something, and you cannot ask anyone about it.*” (CHW, FGD, Johannesburg District). The CHWs also bemoaned the lack of privacy while participating in capacity building workshops: “…*we write pre-tests and post-tests weekly. When we write the tests, we need a quiet space to concentrate, but there will be facility staff members going back and forth while we are writing the test. While we were sitting here, another person would use the microwave.”* (CHW, FGD, Johannesburg District). In the rural sites, the CHWs did not have workspace in the main facility, instead they were accommodated in other buildings, which limited interactions and collaboration with facility-based staff. However, teams with senior supervisors often had access to suitable spaces within the facility, as their supervisors were able to negotiate for them to use the spaces without being interrupted.

#### Uniforms

The CHWs felt obliged to purchase work uniforms, as they wanted the community to recognise them as members of the health facility. To achieve this end, they saved a portion of their stipend to buy a uniform. A CHW commented: “*I also think that the money is little because they still expect us to be presentable when we go to the households. We buy everything that we wear; they don’t offer us anything like a t-shirt for work – nothing*” (CHW, FGD, Tshwane District Team 2). The CHWs felt if they visited the households dressed in private clothes, they will struggle to gain the trust and acceptance of the community. “*It is a must because you have to be presentable to the patients in the community, you cannot go to the community wearing a grey shirt, they will think you are a thief”* (CHW, FGD, Johannesburg).

A facility manager in Tshwane agreed with the CHWs, that a uniform plays a critical role in ensuring the CHWs are recognised as members of the local health facility: “*When someone looks presentable, they gain peoples’ trust and attention easily. As long as the CHWs don’t have uniforms, name tags and continue receiving low stipends, the community will not take them seriously*” (Facility Manager, Interview, Tshwane District Team 4).

#### Relationship with facility-based staff members

Across the different sites, the CHWs reported complex relationships with their supervisors, facility-based staff members and programme coordinators. Due to their low status in the health system, the CHWs felt neither appreciated nor respected by facility-based staff: “*They disrespect and undermine us but we do respect them, not that we are scared of them.” (*CHW, FGD, Johannesburg). The CHW reported that in other facilities some CHWs who felt disrespected by facility staff resorted to physically fighting as a way of asserting themselves (CHW, FGD, Johannesburg District). Despite poor relationships, the CHWs continued to assist the facilities in their daily functions. We observed the Sedibeng teams delivering long-term medications to the elderly, while other teams undertook contact tracing, particularly those defaulting on their long-term medications and made referrals to the facility. However, the CHWs felt facility staff delayed attending to patients the CHW had referred to the facility, and this affected their ability to bring uncooperative patients for care. These issues were better managed in facilities where a senior supervisor was present. In Sedibeng, the senior supervisor negotiated with facility-staff to support the CHWs with timely attendance to referred cases. This arrangement motivated the CHWs and built their work morale.

A facility manager in a Tshwane facility argued there is a need to integrate the programmes into facilities: “*They introduced WBOT as a program as if it is independent from the clinic hence there isn’t unity between WBOT and facility staff. Even worse they employed managers for WBOT, this implies the facility and WBOT are two different entities. So, when they go to the facility manager to ask for medication, the facility refuses and says that they are finishing their stock*” (Facility Manager, Interview, Tshwane District Team 4).

#### Supervision

There were visible differences in how supervision was provided at the different sites. In the urban sites, every morning before going into the community, the supervisor provided the CHWs with in-service training focusing on the cases they attended to the previous day and accompanied them on household visits to provide support. In the rural sites, CHWs were managed by senior supervisors, but there was no morning meeting before going into the community, and they operated without community supervision as their supervisors were not able to go to the field with them, mainly due to the lack of transport.

Infrequent supervision was a concern for the CHW representatives. In Sedibeng, the CHW representatives complained that the supervisors were not investing sufficient time in attending to the needs of the CHWs, instead spent most of their time in the facilities: “*When they get in the clinic, they do clinical work, like children’s immunizations, family planning, or whatever. They don’t support the CHW programme, you understand? It is the challenge that we have, and you find out that when the CHWs are supposed to tell the [CHW representatives] about this, it causes conflict with the supervisors*” (CHW Representative, Interview 2, Sedibeng District). The representatives were unhappy that supervisors worked in the facility while they were employed to oversee CHWs activities in the community. The Department of Health, which is the employer of the supervisors, expect all CHW supervisors to spend 70% of their time in the community, 30% in facilities mainly performing administrative duties related to their functions.

### CHW labour mobilisation

#### Emergence of CHW representation

Prior to joining the nation-wide CHW programme in 2011, the CHWs worked for NGOs providing home-based care services (Table 2). While in the NGOs, the CHWs established a committee to challenge the exploitation perpetuated by the organisations’ management. After joining the national programme, representatives of the CHWs were elected to what became known as the task team. The task team is made up of CHWs, lay counsellors and health promoters working in different facilities within the district. These CHW representatives decided not to become members of established labour unions, as existing unions were perceived as ineffective and with history of siding with management. A member of the task team commented: “*We have tried to be under a labour union, but whenever they left to go to the negotiations, they did not take us. They don’t even take one of us to be a part of it, even if the person is just an observer*” (CHW representative, Interview 1, Sedibeng District).

Another member added: “*The union that we will agree to be under will be a union that we will be a part of. Whenever they go and negotiate for us, or whatever, we must be included and we must be a part of that negotiation*” (CHW representative, Interview 1, Sedibeng District).

The task team in Sedibeng ensured each of the 4 subdistricts of Sedibeng had a representative: “*We have divided ourselves according to clinics; let us say he/she works at facility X. He/she is the one who is responsible for the clinics in closer to her facility. I will also be responsible for facilities closer to where I work. We divide ourselves like that*” (CHW representative, Interview 2, Sedibeng District). Donations from the CHWs funded the travel costs so the task team members could undertake their activities at the different facilities.

In Tshwane and Johannesburg districts, there were emerging groups of CHWs who advocated for the rights of the CHWs. In the rural sites, where the programme was under the management of NGOs, CHWs did not have formal representation; researcher and data collector observations revealed that the CHWs seemed fearful of being outspoken about the need to have a labour union to represent them.

#### Negotiating for decent work

Since 2015, the CHW leaders have led multiple strikes and stay aways in Gauteng province. One strike was triggered by the appointment of the payroll administration company without consulting the CHWs. The protest was directed at the National Department of Health. A member of the task team commented: “*We had a strike and spent about 3 days sleeping there. We came back and went back again to fight and spill rubbish in the offices, but unfortunately, we fought without any luck*” (CHW representative, Interview 2, Sedibeng District). The CHWs were concerned that the department was able to award a multi-million contract but never considered their demands for permanent employment with better pay: “*We wanted to understand why we were tendered without our consent? The department needs our services; why is it outsourcing us when it has money for tenders? That was our concern.*” (CHW representative, Interview 2, Sedibeng District).

The Sedibeng task team leaders were arrested after embarking on this violent, unprotected strike^1^. The task team encouraged the CHWs to withdraw their services and to protest in support of their leaders at the court as their case was being heard. Some CHWs were not keen to participate, as it meant leaving their workplace and neglecting patients. Some CHWs who chose not to participate were threatened with violence. A facility staff member commented: *“They all donated a R10 for transport for those who will go on strike and support the task team. The CHWs who want to work were receiving threats that they will come for them” (*Nurse Mentor, Interview 3, Sedibeng District). Following release of the task team from jail, the CHWs were ordered to withdraw their services in celebration. A facility staff member commented: “*On Wednesday 23th, CHW’s didn’t come to work because they were told by task team to go on a 3 day holiday to celebrate their struggle and past imprisonment of their two task team leaders. There were no formal notification of this holiday and all WBOT leaders, including supervisors didn’t know except [district management]*” (Nurse Mentor, Interview 2, Sedibeng District).

After the unprotected strike action, the task team members continued efforts to improve conditions and access to resources. Within the health facilities, the representatives focused on addressing CHW conflicts with facility staff members and lack of resources: “*We go to the facility and talk to the management and the CHWs about their issues. We request for a space for the CHWs, and if there is no space, the space must be created and they must have the space*” (CHW representative, Interview 2, Sedibeng). One of the accusations was that supervisors neglected the CHWs by not going to the community with them, they are left to fend for themselves while out in the community. The task team also ordered CHWs who were assisting facilities with administrative duties to withdraw their services: **“***She stopped because the task team told her to stop. They must focus on being the CHWs. They are not receptionists*” (Nurse Mentor, Interview 4, Sedibeng District).

The district official responsible for the CHW programme called a meeting aimed at resolving tensions between CHWs and supervisors. At this meeting, the CHW representatives verbally abused the supervisors in the presence of the district and payroll administration officials: “*Some of the [supervisors] were literally crying and some didn’t want to talk. They were busy going up and down, going to the bathroom. They were talking outside. I don’t know what they were saying*” (Nurse Mentor, Interview 3, Sedibeng District). Due to the confrontational approach adopted by the task team, the officials didn’t defend the supervisors or allow the supervisors to speak for themselves. As a result, the supervisors were left exposed at the meeting and the militant approach adopted by the task team created greater hostility between the parties. The nurse mentor reported: “*The task team has more influence than unions or district coordinators because they have control over the CHWs. For example, if they were to call the CHWs today and tell them not to go to work, the CHWs will do exactly that*”.

To resolve the conflicts, at the suggestion of a nurse mentor employed in two of the facilities where we collected data, the task team was invited to a meeting to discuss areas of discomfort with supervisors and to work as partners. This engagement enabled the supervisors to be sensitive to the CHWs’ attempts to seek improved conditions of employment. The task team also changed their approach to dealing with the conflict: “*When there is a concern, the person who complains is called together with the person that they are concerned about. We get together with them in the same room. We listen to the complainant’s story at the same time the defendant is present and is also listening and they are able to respond.*” (CHW representative, Interview 2, Sedibeng District).

In 2018, the Gauteng provincial legislature, an oversight body, initiated a meeting of the CHWs to learn about their challenges in providing community-based care under the national programme. All CHWs from the province were invited to the meeting, those who attended expressed their concerns over their yearly contracts, meagre remuneration, lack of working tools and poor relationships with facility-based staff: “*Mr Motsoaledi [Minister of Health] promised us in November that he is absorbing us. When we were called to Pretoria [National DoH] we thought that we were going to sign contracts as promised but he failed us.”* (CHW, FGD, Tshwane District Team 2). The CHWs expressed their frustration at the lack of progress being made by the DoH to provide permanent employment. The legislature promised to attend to the CHWs grievances and provide feedback.

#### Labour unions

Though the task team held numerous meetings with district management, they had limited negotiating powers, as they could not participate in the Department of Health Bargaining Council (DOHBC). The DOHBC is a formal structure consisting of registered unions, where workers grievances (e.g., need for improved remuneration) are tabled for consideration by the employer. The task team needed representation in the DOHBC to escalate their demands. In 2018, the CHWs in Sedibeng joined National, Education, Health and Allied Workers Union (NEHAWU), a registered labour union, to represent them in the DOHBC. The union was entrusted with negotiating for permanent employment, decent remuneration of the CHWs and other benefits associated with formal employment (e.g. leave allowance). One of the representatives of CHWs was also co-opted into the union regional structures. Some CHWs did not join the union, as the decision appeared rushed, with minimal consultation. Others were concerned that they would not be able to afford the monthly premiums.

#### Outcome of the mobilisations

When fieldwork ended, the task team and union had not been able to secure permanent employment for the CHWs. However, the union had negotiated with the Department of Health to increase the CHW stipend from R2 500 (136 USD) to R3 500 (192 USD). The CHWs felt motivated by the increment: “*It has motivated me because since we got the increase, I have been working harder than before*” (CHW, Interview, Sedibeng District). Another CHW added with the increment they are able to afford necessities and uniform: “*Like now, there is a yellow T-shirt costing about R150, we are able to buy and pay for transport to come to work. On the other side, we buy groceries at home..”* (CHW, Interview, Sedibeng District).

Although the CHWs were excited to receive the increment, they still wanted to be absorbed as permanent workers by the government. The union committed to continue engaging the Department on this issue and other concerns such as leave allowance.

On the facility level, the task team recorded some successes, a member of the task team commented: “*In facility X, the CHWs did not have access to the photocopy machine because they did not contribute to facility budget to buy paper for photocopy machine…how do you expect someone who is being underpaid to have money for such expenses? We fought for them to have access to the printer*” (CHW representative, Interview 2, Sedibeng District).

## DISCUSSION

In the paper, we have reported the CHWs were poorly supervised, resourced and received meagre remuneration, their employment outsourced, without employment benefits and protection. The lack of career progression opportunities demotivated the CHWs, particularly those keen to establish a career in health. In the semi-urban sites, CHWs established a task team to represent them but refused to join a formal union for fear that union would push management insufficiently for their demands. The task team held regular meetings and led protests against clinic, district and provincial management to demand improved conditions of employment. After the recognition by the local provincial legislature, the task team agreed joined a labour union (NEHAWU) to order to be able to participate in the local Bargaining Council. Though they were not successful in getting the government to provide permanent employment, the union negotiated an increase in stipend from R2 500 to R3 500. In contrast, in the rural sites, the CHWs were not actively demanding permanent employment due to their employment contracts being partly managed by NGO managements; they were fearful of being recalled from the government programme.

Since the end of data collection, South African CHWs have engaged in further industrial action. In 2021, NEHAWU took the Department of Health to labour court to declare the 12 months contracts illegal and in contravention of the national labour laws, which state employment cannot be offered on renewable basis for a period exceeding 3 months [19, 20]. The court arbitrator ruled against the union citing a 2018 agreement signed by government and unions. The agreement refers to government dependence on an external grant to pay CHW current salaries, such that CHWs permanent employment is unsustainable.

During the peak of the COVID pandemic, the Gauteng government did not renew the contract of the payroll administration company and offered the CHWs employment contracts similar to other permanently employed government staff members such as nurses [25, 26]. The CHWs monthly salary increased to between R9 000 (495 USD) to R11 000 (605 USD). The COVID-19 pandemic reinforced the need for stable CHW programmes. In many countries, CHW undertook case identification, participated in screening of people with COVID-19 symptoms, traced contacts and encouraged vaccine uptake [21–23]. The CHWs efforts helped stabilise strained healthcare systems, particularly in LMICs. However, poor access to working tools, supportive supervision and decent remuneration limited their effectiveness. A multi-country study conducted in Bangladesh, Pakistan, Sierra Leone, Kenya and Ethiopia showed CHWs were often expected to provide these services without personal protective equipment (PPE), and were unremunerated for the extra tasks they carried out during the pandemic [24]. In India, ASHAs assumed roles of providing antenatal care services because the auxiliary nurses were unavailable [24]. In Brazil, at the height of COVID-19 infections, CHWs protested when they were expected to provide services while being underpaid, poorly trained and with no PPE [18]. The Brazilian CHWs used social media and weekly webinars supported by labour unions to demand safer employment conditions. They took advantage of the vulnerabilities brought by COVID-19 and exercised their agency and capacity to take strategic action [18]. This resulted in some municipalities beginning to purchase PPEs for CHWs undertaking household visits, while some invested in telemedicine to limit CHWs direct contact with patients. As in Brazil, the pandemic in South Africa highlighted the important role that CHWs play in reaching the vulnerable; this helped to advance the CHWs efforts for recognition, particularly in the sites where there was prior mobilisation for improved conditions of employment [25, 26].

As highlighted by the study, in many countries CHWs are without union representation to protect their labour rights [15, 16]. However, following the development of nationwide programmes in different countries, the shift from being disease-specific to comprehensive programmes and the COVID-19 pandemic [13, 24], CHWs have begun to demand better employment conditions [19, 20]. In India, accredited social health activists’ (ASHAs) and anganwadis’^2^ unions have regularly led marches to demand formal employment with benefits [17]. So far, the unions have achieved small gains in terms of increased pay and social security benefits [17].

The study had several strengths and limitations. We spent 3 years collecting data in one of the study sites (Sedibeng district) which allowed us to familiarise ourselves with the history of the programme and conflicts between the CHWs and the Department of Health in the district. In the other districts, data collection was limited to three months. Due to our prolonged stay in the Sedibeng district, familiarity with the CHW programme and labour mobilisation, we used the site as the main comparator to the other sites. Second, the data collection tools were written in English, some participants preferred being interviewed in other local languages (e.g. IsiZulu and Sesotho). To achieve consistency of the translations, as part of pre fieldwork training, data collectors participated in role-plays to practice posing the same question in the different languages. This process allowed the data collectors to be comfortable and consistent in posing the interview questions.

## CONCLUSION

Consistent mobilisation as demonstrated by the urban-based teams enabled the CHWs to successfully negotiate salary increments and advance their call for permanent employment. However, in the rural sites CHWs were less able to join or establish labour representation due to fear of reprisal from NGOs management. In optimising the motivation and performance of CHWs, it is important for the government to prioritise full integration of CHWs into the healthcare system, where they will be afforded their labour rights and support.

## Data Availability

All data supporting our work is provided within the manuscript.

## Acknowledgements

We would like to thank the following individuals for their invaluable contribution: the data collectors, key informants, CHWs and their supervisors, health facility staff members and community representatives. The support of the Sedibeng Health District management, in particular the former district director Mrs. Salamina Hlahane and community-based services coordinator Mrs. Bridget Lefhoedi is highly appreciated.

We would also like to thank these organisations for their generous support: Sedibeng, Johannesburg, Tshwane and Ehlanzeni Health Districts, and the UK Medical Research Council (MRC).

## Abbreviations

ASHA: Accredited Social Health Activist
CHW: Community Health Worker
COSATU: Congress of South African Trade Unions
DOHBC: Department of Health Bargaining Council
FGD: Focus Group Discussion
OTL: Outreach Team Leader
PHC: Primary Health Care
PPE: Personal Protective Equipment
LMIC: Low and Middle-Income Country
NEHAWU: National, Education, Health and Allied Workers Union
NGO: Non-Government Organisation
UHC: Universal Health Coverage
WBOT: Ward based Outreach Team
WHO: World Health Organisation

## Footnotes

1 Non-procedural, or unprotected strike, is one where the strikers have not complied with the requirements of the South African Labour Relations Act of 1995 before going on strike. This removes the employer’s opportunity to develop contingency plans for running the business during the strike.

2 These workers care for the health and wellbeing of women, children and other socioeconomically deprived groups.

## References

1. Woldie, M., et al., Community health volunteers could help improve access to and use of essential health services by communities in LMICs: an umbrella review. Health Policy Planning, 2018. 33(10): p. 1128–1143.

2. Ballard, M. and P. Montgomery, Systematic review of interventions for improving the performance of community health workers in low-income and middle-income countries. BMJ Open, 2017. 7(10): p. e014216.

3. Kok, M.C., H. Ormel, and J.E.W. Broerse, Optimising the benefits of community health workers’ unique position between communities and the health sector: A comparative analysis of factors shaping relationships in four countries. 2017. 12(11): p. 1404–1432.

4. Gaziano, T.A., et al., Hypertension education and adherence in South Africa: a cost-effectiveness analysis of community health workers. BMC Public Health, 2014. 14: p. 240.

5. Kok, M.C., et al., Which intervention design factors influence performance of community health workers in low-and middle-income countries? A systematic review. Health Policy and Planning, 2015. 30(9): p. 1207–27.

6. Cometto, G., et al., Health policy and system support to optimise community health worker programmes: an abridged WHO guideline. The Lancet Global Health, 2018. 6(12): p. e1397–e1404.

7. Pandya, S., et al., Understanding factors that support community health worker motivation, job satisfaction, and performance in three Ugandan districts: opportunities for strengthening Uganda’s community health worker program. International Journal of Health Policy and Management, 2022. 11(12): p. 2886–2894.

8. Ormel, H., et al., Salaried and voluntary community health workers: exploring how incentives and expectation gaps influence motivation. BMC Human Resources for Health, 2019. 17(1): p. 59.

9. Maree, J., The emergence, struggles and achievements of black trade unions in South Africa from 1973 to 1984. Labour, Capital and Society/Travail, capital et société, 1985: p. 278–303.

10. Dibben, P., G. Wood, and K. Mellahi, Is social movement unionism still relevant? The case of the South African federation COSATU. Industrial Relations Journal, 2012. 43(6): p. 494–510.

11. Masiya, T., Social movement trade unionism: Case of the congress of South African trade unions. Politikon, 2014. 41(3): p. 443–460.

12. South African Department of Health., Provincial guideliness for the implementation of the three streams of PHC Re-engineering. Government Press, 2011.

13. Schneider, H., et al., The challenges of reshaping disease specific and care oriented community based services towards comprehensive goals: a situation appraisal in the Western Cape Province, South Africa. BMC Health Services Research, 2015. 15: p. 436.

14. Braun, V. and V. Clarke, Thematic analysis. 2012.

15. van de Ruit, C. and A. Breckenridge, South African community health workers’ pursuit of occupational security. Gender, Work & Organization, 2022.

16. Trafford, Z., A. Swartz, and C.J. Colvin, “Contract to Volunteer”: South African Community Health Worker Mobilization for Better Labor Protection. New Solution, 2018. 27(4): p. 648–666.

17. Sinha, D., India community health workers strugge for recognition, in Roar. 2021.

18. Lotta, G. and J. Nunes, COVID-19 and health promotion in Brazil: community health workers between vulnerability and resistance. Global Health Promotion, 2022. 29(1): p. 14–22.

19. Maverick, D., Community Healthcare Workers demand wage increase and recognition as public servants. 2019.

20. Daily Maverick., Arbitrator rules against Nehawu over bid to make contract workers permanent state employees. 2021.

21. Bhaumik, S., et al., Community health workers for pandemic response: a rapid evidence synthesis. BMJ Global Health, 2020. 5(6): p. e002769.

22. Olateju, Z., et al., Community health workers experiences and perceptions of working during the COVID-19 pandemic in Lagos, Nigeria—A qualitative study. Plos one, 2022. 17(3): p. e0265092.

23. Ballard, M., et al., Prioritising the role of community health workers in the COVID-19 response. BMJ Global Health, 2020. 5(6): p. e002550.

24. Salve, S. and J. Raven, Community health workers and Covid-19: Cross-country evidence on their roles, experiences, challenges and adaptive strategies. 2023. 3(1): p. e0001447.

25. Karibu, Community Health Workers to sign permanent contracts. 2020, Karibu Online.

26. Nicole, L., Why are community health workers ditching unions?. 2023, BusinessLive.

